# The Brain Reacting to COVID-19: Analysis of the Cerebrospinal Fluid and Serum Proteome,Transcriptome and Inflammatory Proteins

**DOI:** 10.1101/2022.04.10.22273673

**Authors:** Dirk Reinhold, Vadim Farztdinov, Yan Yan, Christian Meisel, Henrik Sadlowski, Joachim Kühn, Frank H. Perschel, Mathias Endres, Emrah Düzel, Stefan Vielhaber, Karina Guttek, Alexander Goihl, Morten Venø, Bianca Teegen, Winfried Stöcker, Paula Stubbemann, Florian Kurth, Leif E. Sander, Markus Ralser, Carolin Otto, Simon Streit, Sven Jarius, Klemens Ruprecht, Helena Radbruch, Jørgen Kjems, Michael Mülleder, Frank Heppner, Peter Körtvelyessy

## Abstract

Patients with COVID-19 can have a variety of neurological symptoms, but the pathomechanism of CNS involvement in COVD-19 remains unclear. While routine cerebrospinal fluid (CSF) analyses in patients with neurological manifestations of COVID-19 generally show no or only mild inflammation, more detailed data on inflammatory mediators in the CSF of patients with COVID-19 are scarce.

Here, we used mass spectrometry to study the proteome, Enzym-linkend immunoassays, semiquantitative cytokine arrays, autoantibody screening, and RNA profiling to study the neuroinflammation. We study the inflammatory response in paired CSF and serum samples of patients with COVID-19 (n=38). Patients with herpes simplex virus encephalitis (HSVE, n=10) and patients with non-inflammatory, non-neurodegenerative neurological diseases (n=28) served as controls. Proteomics on single protein level and subsequent pathway analysis showed similar yet strongly attenuated inflammatory changes in the CSF of COVID-19 patients compared to HSVE patients. CSF/serum indices of interleukin-6, interleukin-16 and CXCL10 together point at an origin from these inflammatory proteins from outside the central nervous system. When stratifying COVID-19 patients into those with and without bacterial superinfection as indicated by elevated procalcitonin levels, inflammatory markers were significantly higher in those with concomitant bacterial superinfection. RNA sequencing in the CSF revealed 101 linear RNAs comprising messenger RNAs, micro RNAs and t-RNA fragments being significantly differentially expressed in COVID-19 than in HSVE or controls.

Our findings may explain the absence of signs of intrathecal inflammation upon routine CSF testing despite the presence of SARS-CoV2 infection-associated neurological symptoms. The relevance of blood-derived mediators of inflammation in the CSF for neurological post-COVID-19 symptoms deserves further investigation.

## INTRODUCTION

Infections with the severe acute respiratory syndrome coronavirus 2 (SARS-CoV-2) can result in a severe infectious syndrome requiring hospital admission and sometimes mechanical ventilation, which is sometimes accompanied by neurological manifestations such as headache, dys-/anosmia, encephalopathy, and stroke *(1–4)*. However, evidence for direct brain damage due to SARS-CoV-2 infection is scarce and remains controversial *(5, 6)*. Recently a preprint was published claiming that in ACE2 mice, respiratory infection with SARS-CoV-2 is associated with damage to myelin and oligodendrocytes *(7)*.

One major question is whether the neurological symptoms associated with coronavirus disease 2019 (COVID-19) are an indirect sequala of systemic SARS-CoV-2 infection, with the resulting cytokine storm reaching the CNS by passive diffusion or a leaky blood-brain barrier, or by a genuine intrathecal inflammatory response.

Until now SARS-CoV-2 has been detected with high resolution microscopy only in the olfactory bulb and the peri-vascular regions in the medulla oblongata with known leaky blood/brain-barrier *(5, 8, 9)*. Interestingly, microglial activation is prominent in most post-mortem studies of COVID-19 patients *(5, 9)*, pointing at an at least passive reaction of the innate immunological system of the brain *(8–10)*. Moreover invading T cells in autopsy studies on patients with severe COVID-19 were described with a dominant activated effector phenotype from outside the CNS *(11)*. On the other hand, except for evidence of mild to moderate blood-CSF barrier dysfunction, routine parameters in the CSF of patients with COVID-19 with or without additional neurological symptoms did not reveal any general signs of inflammation in the majority of patients *(2, 3, 13, 14)*.

Notably, the studies conducted so far did not distinguish between COVID-19 patients with and without a bacterial superinfection (BSI). However, these superinfections, which occur most often in severely affected patients with COVID-19, may also trigger an immunological response in the CNS biasing the immunological results. We therefore used procalcitonin levels above 1 [pg/ml] as an indicator for bacterial superinfection. Here, we used mass spectrometry, ELISA, semiquantitative cytokine arrays autoantibody screening, and RNA profling to characterize inflammatory responses in the CSF and serum of patients with COVID-19 stratified according to the presence or absence of signs of bacterial superinfection. We also used Proteome analysis of the CSF from COVID-19 patients to elucidate the extent of the general neurological and neuroimmunological pathways and examined proteins of the brain providing a comprehensive overview on differentially expressed proteins. Then, we used RNA sequencing to investigate the profile of linear and circular RNAs in the CSF.

Our results point towards a mild but distinct response of the brain to peripheral inflammation rather than to an autochthonous intrathecal anti-viral immune response. Proteomic analyses showed a significant difference between COVID-19 patients with and without bacterial superinfections. By RNA sequencing, we found linear RNAs and circular RNAs that are deregulated in CSF of COVID-19 patients compared to healthy controls and patients with other neurological diseases.

## RESULTS

### Routine parameters in the COVID-19 cohort showed an almost normal pattern

We studied CSF/serum pairs of a total of 38 patients with COVID-19. Ten patients with HSVE and 28 patients with non-inflammatory and non-neurodegenerative diseases served as controls. Demographic findings and the CSF parameters of patients and controls are summarized in Table 1. Consistent with previous findings, routine CSF parameters of patients with COVID-19 showed normal levels with only the albumin CSF/serum ratio (median 10.24, range 5-43.6) being slightly higher than normal (Tab1).

**Table 1.**
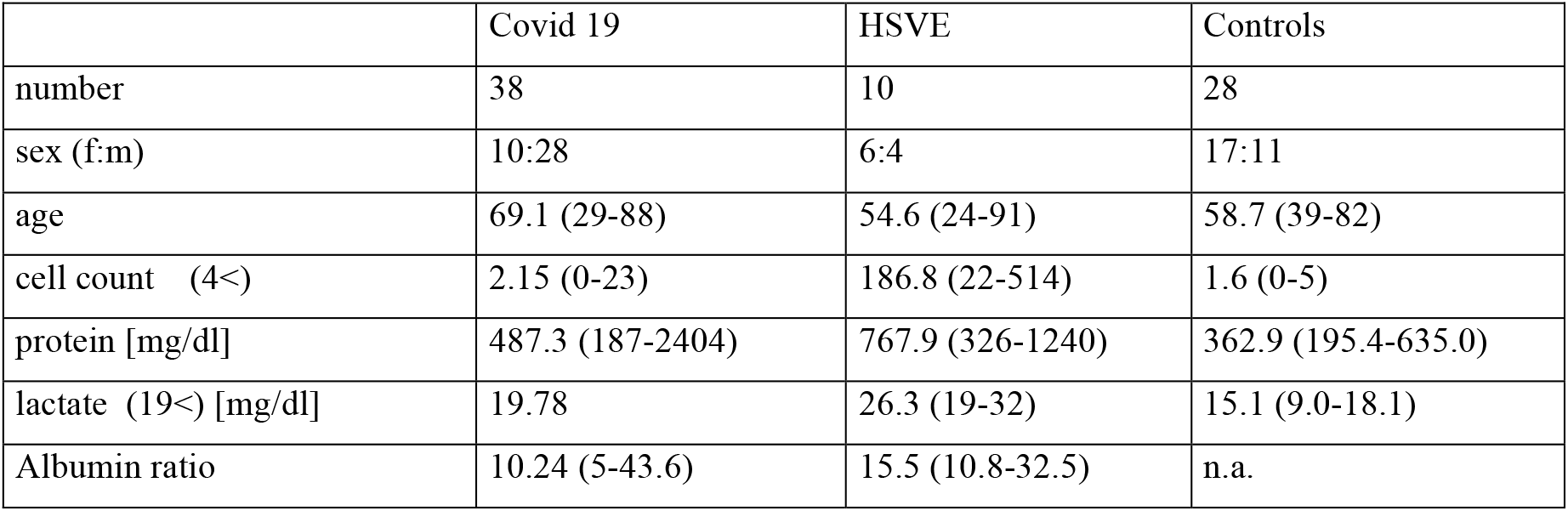
Epidemiological data and routine CSF results from the entire COVID-19 cohort, the HSVE cohort, and the proteomics controls. Note that control groups in the RNA measurements are different and described in the Methods section.

|  | Covid 19 | HSVE | Controls |
| --- | --- | --- | --- |
| number | 38 | 10 | 28 |
| sex (f:m) | 10:28 | 6:4 | 17:11 |
| age | 69.1 (29-88) | 54.6 (24-91) | 58.7 (39-82) |
| cell count (4<) | 2.15 (0-23) | 186.8 (22-514) | 1.6 (0-5) |
| protein [mg/dl] | 487.3 (187-2404) | 767.9 (326-1240) | 362.9 (195.4-635.0) |
| lactate (19<) [mg/dl] | 19.78 | 26.3 (19-32) | 15.1 (9.0-18.1) |
| Albumin ratio | 10.24 (5-43.6) | 15.5 (10.8-32.5) | n.a. |

### CSF proteomics show an almost similar but attenuated inflammatory response in patients with COVID-19 as compared to HSVE

We used mass spectrometry to comprehensively analyze the CSF proteome in patients with COVID-19 as compared to patients with HSVE. Comparing the activated proteins in COVID-19 and HSVE show almost similar activation pattern but differences in both protein level and regulation as seen with the direct comparison of regulated proteins in both cohorts (figure 1A, contrasts III and IV, see tab 2 for contrast overview). Several proteins are regulated in response to COVID-19 either stronger or in different directions than in HSVE. We found a large group of immunoglobulins and most of complement components among proteins showing a similar but very attenuated response in COVID-19. Proteomics revealed a downregulation (in opposite to HSVE direction) of apolipoproteins APOA1, APOA2, APOA4, APOC1; also an equal or stronger downregulation of CACNA2D1, NPTXR, NRCAM, NRXN2, SEMA7A, SIRPA, SLITRK1, VGF proteins (full list of protein abbreviations Supplement Table 1). On the other hand, we could detect a stronger upregulation in COVID-19 of complement proteins C9, CFD, fibrinogen proteins FGA, FGB, FGG and some other proteins (AMBP, ITIH4, leucine-rich alpha-2-glycoprotein LRG1, alpha-1-acid glycoproteins ORM1, ORM2, SERPINA1, SERPINA3, SERPINF2, SERPING1, SGCE, PROCR, RBP4). A different protein upregulation than HSVE was detected for insulin-like growth factor-binding proteins IGFBP2, IGFBP4, IGFBP6.

**Figure 1.**
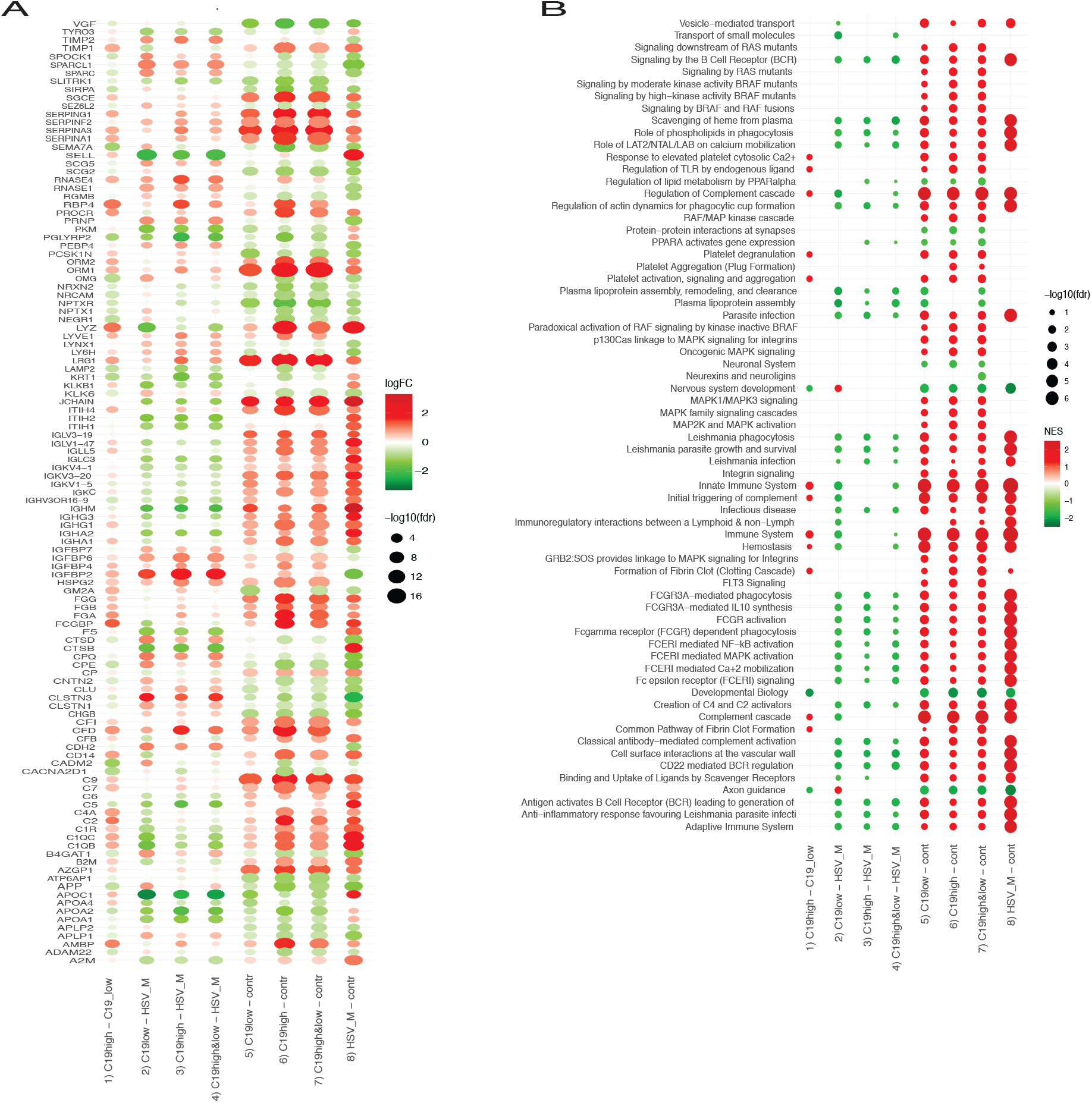
Proteomics analysis. **(A)** Dot heatmap of significantly regulated proteins across all contrasts. The proteins satisfy selection criteria: alpha= 0.001 and |logFC| > 1 at least in one of the eight contrasts (see Table 1), here shown as x-axis labels. All contrasts in this and the next plot are split into four groups: group I for comparison of C19 high PCT vs. C19 low PCT, group II for comparison of C19 groups vs. HSVE, group III for comparison of C19 groups vs. controls and group IV for comparison of HSVE group vs. controls. One can see that the strongest regulation (by the number of proteins with high |logFC|) is observed for HSVE vs. control (group IV). Regulation in group of contrasts III (COVID-19 groups vs. control, contrasts 5 – 7 in Table 1) appears as following the regulation in group IV (contrast 8 in Table X2). Still, the extent of the regulation is different, as can be seen from the pattern of protein regulation in group of contrasts II (COVID-19 groups vs. HSVE, contrasts 2-4 in Table 1). **(B)** Dot heatmap of significantly enriched REACTOME terms across all contrasts. Results of GSEA using REACTOME functional terms with fdr<0.05 at least in one of the eight contrasts (see Table 1). All contrasts in the plot are split into four groups: group I for comparison of C19 high PCT vs. C19 low PCT, group II for comparison of C19 groups vs. HSVE, group III for comparison of C19 groups vs. controls and group IV for comparison of HSVE group vs. controls.

**Table 2.** Contrasts. addressed in the Model 1 analysis. C19 = COVID-19, C19H = COVID-19 with high PCT, C19L = COVID-19 with low PCT, HSVE = HSV meningoencephalitis.

| Contrast | Description | Short Notation |
| --- | --- | --- |
| Contrast 1 | C19_high_PCT vs. C19_low_PCT | C19H/C19L |
| Contrast 2 | C19_low_PCT vs. HSV Meningoencephalitis | C19L/HSVE |
| Contrast 3 | C19_high_PCT vs. HSV Meningoencephalitis | C19H/HSVE |
| Contrast 4 | (C19_low_PCT+C19_high_PCT)/2 vs. HSV meningoencephalitis | C19H&L/HSVE |
| Contrast 5 | C19_low_PCT vs. control | C19L/cont |
| Contrast 6 | C19_high_PCT vs. control | C19H/cont |
| Contrast 7 | (C19_low_PCT+C19_high_PCT)/2 vs. control | C19H&L/cont |
| Contrast 8 | HSV meningoencephalitis vs. control | HSVE/cont |

### Bacterial superinfection, as inferred from high PCT levels, aggravates the host response

Many proteins and the response protein pattern in COVID-19 patients with BSI and in COVID-19 patients without BSI (Covid19_highPCT_ vs. Covid19_lowPCT_) are differently regulated compared to controls (Covid19_highPCT_ + Covid19_lowPCT_ vs. controls). Similarly, the response protein pattern in COVID-19 patients with BSI (Covid19_highPCT_ vs. controls) is highly correlated (*r* = 0.94) to the response in COVID-19 patients without BSI (Covid19_lowPCT_ vs. controls).

When analyzing the response of proteins in COVID-19 patients with BSI compared to COVID-19 patients without BSI some proteins show monotonic increase of response with the increase of log_2_(1 + *PCT*). Combined log2 expression of the three proteins C4a, CD14 and NRCAM (*y*= (*C4A* + *CD14*)/2 – *NRCAM)* used in a simple linear fit conducted on all COVID 19 samples resulted in a dependence with rather high coefficient of determination, R^2^ = 0.67 (see supplementary figure 1), meaning that these proteins and their pathways are only activated in COVID-19 with BSI.

The comparisons of CSF proteomics from HSVE patients with controls show an overwhelming activation at almost every level in HSVE patients (fig1B). The COVID-19 cohort comparison to the same controls are similar as the HSVE pattern of regulation but on a much lower level. In total, more pathways were activated in the CSF proteomics of COVID-19 patients as in HSVE patients. The pathways connected with lipoproteins and nervous system development are especially enriched and downregulated, whereas the pathways connected with integrins, and fibrin clot formation are enriched and upregulated.

### Cytokine indices reveal no intrathecal production of cytokines

For a general assessment of the inflammatory response 36 cytokines were measured in the CSF samples of COVID-19 (n=9), HSVE (n=5) and non-inflammatory, non-neurodegenerative controls (n=4) with a semiquantitative array technique via a background-corrected sum intensity analysis as described before *(15)*. The analysis yielded a plethora of different elevated cytokine levels compared to controls (figure 2). Mean levels of CCL1, CXCL10, IL-1ra, IL-6, IL-8 and IL-16 as well as Serpin E1 and C5/C5a are elevated in the CSF of COVID-19 patients compared to non-neurological controls. Patients suffering from the severe neuroinflammatory disease HSVE had in general higher cytokine levels then any other disease. Only the level of IL-16 was in general higher in COVID-19 patients CSF than in HSVE patients with every other cytokine level being higher in the CSF of HSVE patients. We therefore excluded HSVE in general from statistics and used it only as a reference for cytokine ratios and proteomics in neuroinflammatory diseases.

**Figure 2.**
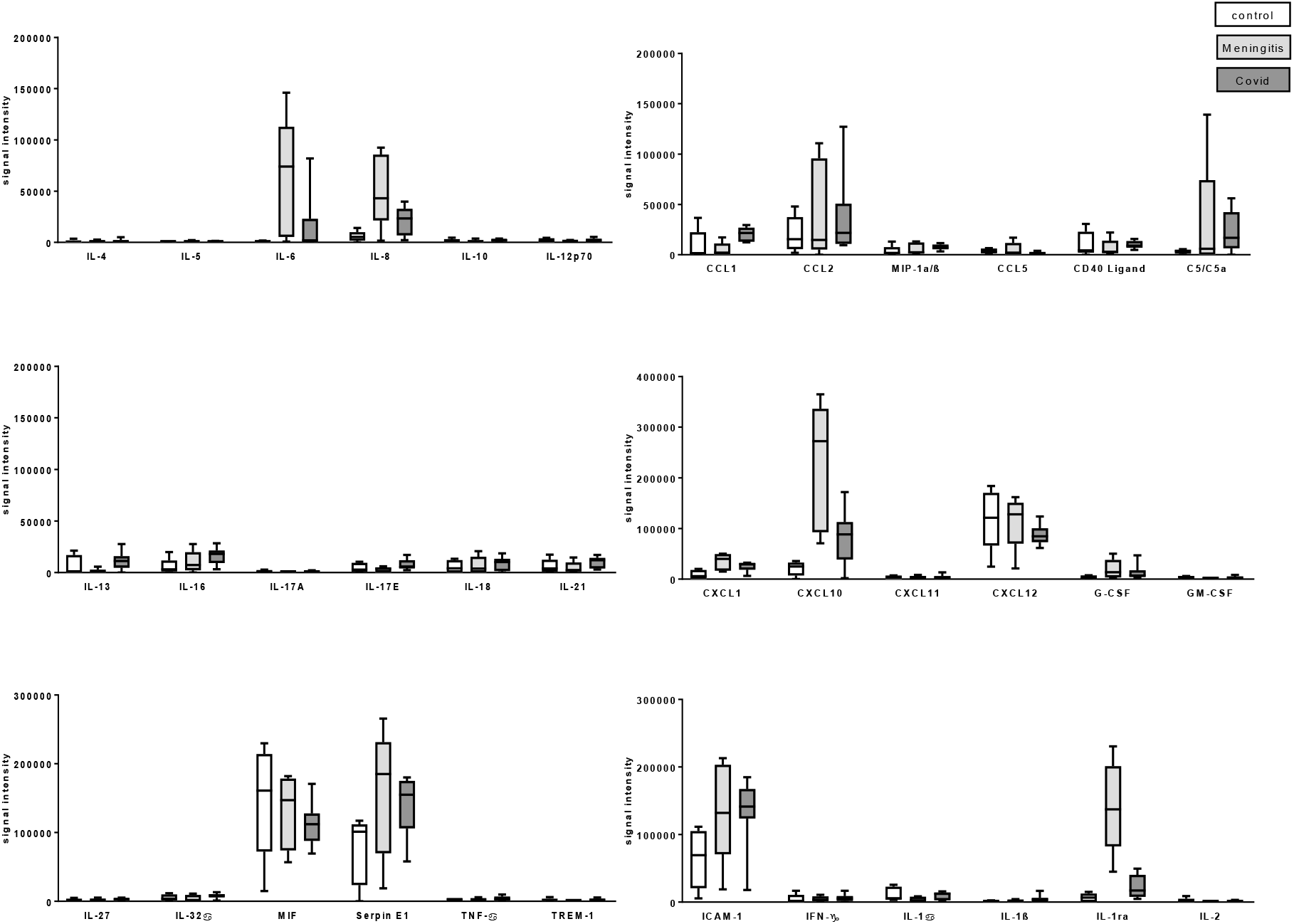
Results of the semi-quantative cytokine arrays as box-plot.

We further examined candidate cytokines with specific ELISA (see tab 3 and 4 and figure 3). This enabled us to calculate ratios between CSF and serum to determine the origin of each measured cytokine. Cerebrospinal fluid/serum ratios under 1 show indicate a higher serum level. Non-neurological controls showed mean IL-6 concentrations of 3.2 pg/ml (range 0.5-9.7 pg/ml) in the CSF and 2.6 pg/ml (range 0-9.4 pg/ml) in the serum resulting into a CSF/serum IL-6 ratio of 1.2. Mean IL-6 levels as in the CSF of C19_total_ patients was 37.0 pg/ml (range 0.7-552.1 pg/ml) and in serum 92.5 pg/ml (range 0.7-552.43 pg/ml) (tab 2 and Fig3A). Every IL-6 CSF/serum ratio in every COVID-19 patient was between 0.15-0.55 indicating an IL-6 origin from blood invading the CSF compartment. Patients with COVID-19 and low PCT had much lower mean IL-6 levels in the CSF (7.0 pg/ml, (range 0.7 pg/ml – 24.7 pg/ml)) and serum (51.3 pg/ml (0.75 pg/ml - 345.5 pg/ml)) than patients with COVID-19 and high PCT (serum 174.9 pg/ml (7.1-552.4 pg/ml) and CSF with mean 96.9 pg/ml (range 1.5-552.1 pg/ml). Serum IL-6 levels in the C19_highPCT_ patients compared to our non-neurological cohort were significantly higher (p<0.01). Significant differences (p<0.05) between IL-6 levels in the C19_highPCT_ cohort and the C19_lowPCT_ cohort in the CSF were found but not between serum IL-6 levels, nor ratios. In sum, CSF IL-6 levels are very likely driven by the serum IL-6 levels in COVID-19 patients. These results are in contrast to HSVE patients with a mean CSF IL-6 concentration of 315.5 pg/ml (range 1.77-552.9 pg/ml) and in serum of 12.7 pg/ml (range 0.3-573.3 pg/ml) resulting in a mean ratio of 24.84, pointing at the CNS as IL-6 source (see table 4).

**Table 3.** Cytokine and progranulin levels and ratios in our groups. IL-6 = interleukin-6, IL-16 = interleukin-16, CXCL10 = C-X-C motif chemokine 10; n.a.= not available, PGRN = progranulin. NC= non-inflammatory, non-neurodegenerative controls

|  | N | Mean<br>serum<br>level<br>(pg/mg) | Range (pg/ml) | Mean<br>CSF level<br>(pg/ml) | Range (pg/ml) | Ratio | Range |
| --- | --- | --- | --- | --- | --- | --- | --- |
| ***IL-6*** |  |  |  |  |  |  |  |
| NC | 11 | 2.6 | 0.0-9.4 | 3.2 | 0.5-9.7 | 1.23 | 0.0-26.8 |
| HSVE | 10 | 12.7 | 0.0-57.35 | 315.5 | 1.77-552.9 | 24.84 | 0.0-57.35 |
| C19 total | 18 | 92.5 | 0.75-552.4 | 37.0 | 0.68-552.1 | 0.4 | 0.02-5.09 |
| C19 lowpct | 12 | 51.3 | 0.75-345.5 | 7.00 | 0.68-24.72 | 0.14 | 0.02-5.09 |
| C19 highpct | 6 | 174.9 | 7.1-552.4 | 96.9 | 1.54-552.1 | 0.55 | 0.02-0.98 |
| ***IL-16*** |  |  |  |  |  |  |  |
| NC | 11 | 206.1 | 165.3-275.8 | 45.2 | 39.3-57.6 | 0.22 | 0.16-0.3 |
| HSVE | 5 | 253.2 | 209.8-316.6 | 115.7 | 73.8-18.8 | 0.46 | 0.31-0.53 |
| C19 total | 18 | 358.9 | 109.7-775.8 | 53.3 | 44.7-74.8 | 0.15 | 0.09-0.52 |
| C19 lowpct | 12 | 295.0 | 109.7-521.2 | 52.4 | 44.7-67.2 | 0.18 | 0.11-0.52 |
| C19 highpct | 6 | 486.8 | 226.2-775.8 | 55.6 | 43.3-74.8 | 0.11 | 0.08-0.19 |
| ***CXCL10*** |  |  |  |  |  |  |  |
| NC | 11 | 161.6 | 91.37-368.4 | 205.4 | 89.26-416.1 | 1.27 | 0.6-4.04 |
| HSVE | 10 | 256.5 | 83.37-627.5 | 894.8 | 285.9-1087.3 | 3.49 | 1.56-13.04 |
| C19 total | 18 | 405.2 | 32.77-1234.3 | 226.7 | 49.06-622.1 | 0.56 | 0.08-7.71 |
| C19 lowpct | 12 | 301.8 | 32.77-765.7 | 178.0 | 13.91-622.1 | 0.59 | 0.08-7.71 |
| C19 highpct | 6 | 612.1 | 57.92 | 324.0 | 49.83-565.1 | 0.53 | 0.2-0.98 |
| ***PGRN*** |  |  |  |  |  |  |  |
| NC | 22 | N.a. | N.a. | 0.9963 | 0.49-2.09 | N.a. | N.a. |
| MHSVE | 6 | N.a. | N.a. | 5.6717 | 1.04-9.81 | N.a. | N.a. |
| C19 total | 23 | N.a. | N.a. | 1.675 | 0.67-3.04 | N.a. | N.a. |
| C19 lowpct | 13 | N.a. | N.a. | 1.678 | 0.67-3.04 | N.a. | N.a. |
| C19 highpct | 10 | N.a. | N.a. | 1.671 | 0.75-2.22 | N.a. | N.a. |

**Table 4.** Mann-Whitney U-test on IL-6, Il-16, CXCl10. Note that progranulin is also synthesized outside the CNS and therefore serum values and ratios were not calculated. *n*.*a*.*= not available*. NC= non-inflammatory, non-neurodegenerative controls

|  |  | **Progranulin** | **IL6** | **IL16** | **CXCL10** |
| --- | --- | --- | --- | --- | --- |
| **CSF** | NC vs C19 total | <0.001 | 0.4 | <0.001 | 0.76 |
|  | NC vs C19 low pct | <0.001 | 0.09 | <0.001 | 0.65 |
|  | NC vs C19 high pct | <0.001 | 0.17 | <0.001 | 0.2 |
|  | C19 low pct vs<br>C19 high pct | 0.98 | 0.17 | 0.49 | 0.1866 |
| **Serum** | NC vs C19 total | n.a | 0.07 | <0.001 | <0.001 |
|  | NC vs C19 low pct | n.a | 0.13 | <0.001 | 0.12 |
|  | NC vs C19 high pct | n.a | <0.001 | <0.001 | <0.001 |
|  | C19 low pct vs<br>C19 high pct |  | 0.12 | <0.001 | 0,09 |
| **CSF/**<br>**serum**<br>**ratio** | NC vs C19 total | n.a | 0.12 | 0.18 | 0,56 |
|  | NC vs C19 low pct | n.a | 0.25 | 0.62 | 0,92 |
|  | NC vs C19 high pct | n.a | 0.27 | <0.001 | <0.001 |
|  | C19 low pct vs<br>C19 high pct |  | 0.23 | 0.08 | 0,37 |

**Figure 3.**
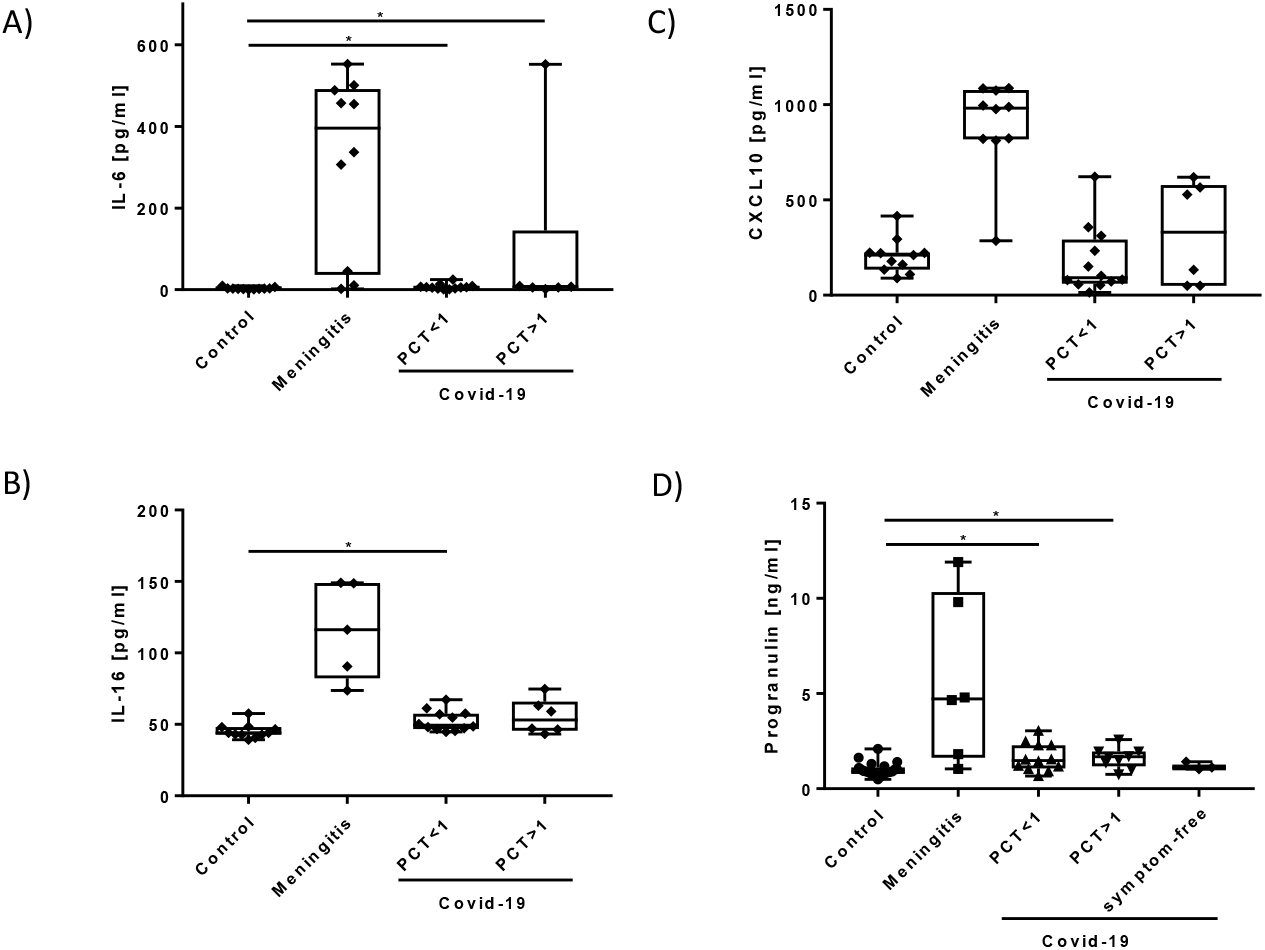
Cytokine and progranulin levels in the CSF. Please note that HSVE levels were not part of the statistical analysis due to the high levels of cytokines. (A) Interleukin-6 (IL-6). (B) Interleukin-16 (IL-16). (C) CXCL10. (D) Progranulin levels. * p<0.05; ** p<0.01

IL-16 levels in CSF and serum were significantly higher in every COVID-19 patient than in non-neurological controls (see table 4 for results and figure 4B). Significant differences were found in IL-16 concentrations in the serum between the C19_lowPCT_ cohort and C19_highPCT_ cohort (p<0.02) but not in CSF (p=0.48) with serum IL-16 levels in the C19_lowPCT_ cohort reaching a mean = 295 pg/ml (range 109.7 - 521.2 pg/ml) and in the C19_highPCT_ cohort with a mean = 486.6 pg/ml (ranging from 226.2 - 775.8 pg/mg). Every CSF/serum ratio for IL-16 was between 0.11- 0.46 showing an origin from outside the CNS. This ratio turned significant when comparing IL- 16 ratios from non-neurological controls with a highly inflammatory condition such as the C19_highPCT_ cohort (p< 0.01).

**Figure 4.**
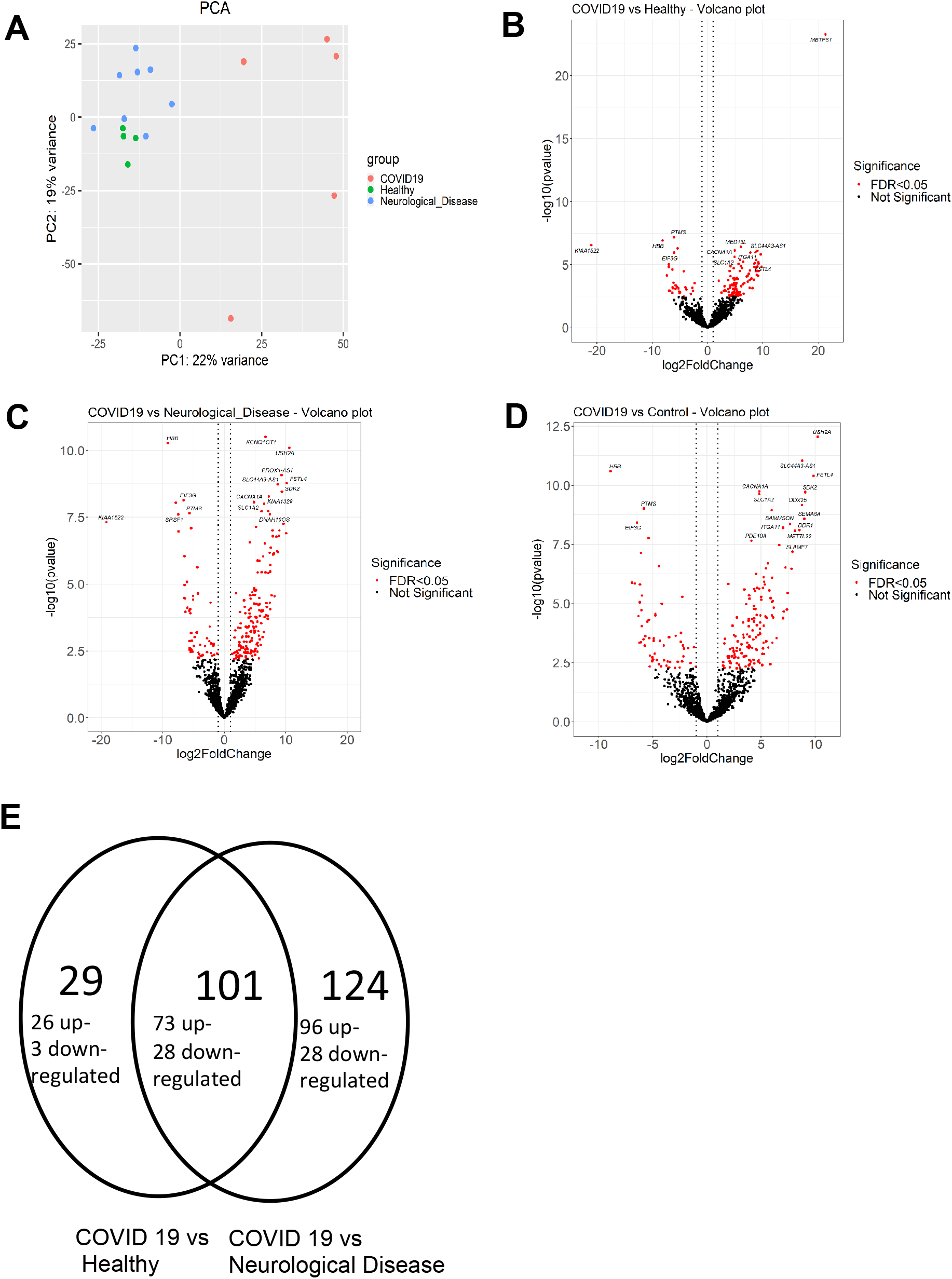
Linear RNA data analysis. (A) Principal component analysis (PCA) of linear RNA profile in the CSF of COVID-19 patients, healthy controls and patients with neurological disease (AD and MS). Differential expression analysis of linear RNAs in (B) CSF of COVID-19 patients and healthy controls, (C) CSF of COVID-19 patients and patients with neurological disease and (D) CSF of COVID-19 patients and all controls. Red dots: FDR < 0.05.

CXCL10 levels in serum of the C19_Total_ cohort (serum= 405.2 pg/ml, range 32.8 pg/ml - 1234.3 pg/ml) are significantly higher (p< 0.05) as in non-neurological controls (Serum = 161.6 pg/mg, range 91.4 – 368.4 pg/ml) (figure 4C). In general, CXCL10 levels in the serum were higher than in the CSF for the entire COVID-19 cohort (serum= 405.2 pg/ml, 32.8 - 1234.3 pg/ml versus CSF = 226.7 pg/mg, 49.1 - 622.1 pg/ml) resulting in CSF/serum ratios in between 0.53-0.59 pointing at a cytokine origin from outside of the CNS.

### Progranulin levels point to microglial activation or lesions in the olfactory system

Progranulin is an anti-inflammatory protein produced by activated microglia and neurons throughout the brain with the highest neuronal concentrations detected in the olfactory and temporal regions of the brain *(16–18)*. The mean progranulin concentration in the total COVID-19 cohort was 1.68 ng/ml (range 0.67 - 3.04 ng/ml). This was significantly higher than in non-inflammatory, non-neurodegenerative controls (mean 1.00 ng/ml, range 0.49-2.09 ng/ml) (p<0.01) (figure 4D).

No significant difference in progranulin levels was found between the C19_lowPCT_ (mean 1.68 ng/ml, range 0.67-3.04 ng/ml) subgroup and the C19_highPCT_ subgroup (mean 1.67 ng/ml, range 0.75-2.58 ng/ml). Three asymptomatic COVID-19 patients in whom the infection with SARS-CoV2 was accidently found at hospital admission had normal progranulin levels in the CSF (mean 1.19 ng/ml, range 1.03-1.42 ng/ml). Herpes-simplex Virus meningoencephalitis patients had the highest progranulin CSF levels (5.67 ng/ml, range 1.04-11.91 ng/ml) significantly higher than any other cohort.

### Low-titre anti-neuronal antibodies in some patients but no evidence for autoimmune encephalitis

Three out of 32 patients were positive for serum anti-myelin IgG antibodies in a tissue-based assay. As a limitation, such antibodies have been observed also in other diseases and even in healthy controls and may thus be unspecific. In two patients, serum IgG antibodies against contactin-associated protein 2 (Caspr2)-IgG was found at a very low titer (1:32) and very low serum IgG antibodies against glycine receptors (1:10). However, and none of these patients had neurological symptoms typically seen in patients with Caspr2- or glycine receptor-related autoimmunity and no confirmatory tests were performed. One patient with up-beat nystagmus after resuscitation had reportedly a 1:100 titer of NMDA receptor IgG antibodies in the serum. However, CSF was negative, which is atypical for NMDAR encephalitis. and no corresponding IgG binding pattern on native primate hippocampus and cerebellum sections was found. This patient died from hypoxic encephalopathy. Another patient with myoclonia without signs of epilepsy on EEG was positive for serum anti-Yo IgG antibodies in an immunoblot assay at a titre of 1:100 but the result could not be confirmed in by immunohistochemistry, suggesting a false-positive result. Confirmatory testing by means of a CDR2/CDR2L-spefific cell-based assay was not performed. At autopsy, histopathology revealed no signs of cerebellar pathology including normal Purkinje cells.

In total, cerebrospinal fluid/serum pairs from 16/32 patients were screened on native primate hippocampus and rat cerebellum tissue without any pathological results (see supplementary table2). Thus, no pathological auto-antibody results could be seen except one out of 32+32 samples.

### CSF RNA sequencing reveals distinct RNA profiles in COVID-19

RNA profiling of the CSF has been performed to assess another stage of possible neuroinflammatory response. We mapped the reads in the data analysis of RNA sequencing in an exploratory study to both back-splicing-junctions (BSJ) to quantify circular RNAs (circRNAs) and to the linear RNA database to quantify the linear RNAs. In the PCA plot, CSF from patients with a severe COVID-19 (n=5) had a linear RNA profile (figure 4A) and a different circRNA profile (supplementary figure 2) compared to healthy controls (n=4) and neurological diseases (n=8)(ND, comprising 4 Alzheimer Disease patients and 4 Multiple sclerosis patients at diagnosis) We performed a differential expression analysis on the circRNAs and linear RNAs, and the RNAs were considered to be differentially expressed with FDR < 0.05. There were two circRNAs (circ_00022_SMC1A and circ_00007_MAN1A2) downregulated in COVID-19 CSF.

In the linear RNA profiling data set, 169 linear RNAs were up-regulated and 56 were down-regulated in COVID-19 CSF compared to the CSF of ND patients, while 99 linear RNAs were up-regulated and 31 were down-regulated in COVID-19 CSF compared to healthy controls (figure 4 B,C,D). There were 73 linear RNAs up-regulated and 28 linear RNAs down-regulated in COVID-19 CSF compared to the CSF from both ND patients and healthy controls (figure 4D) and the 101 RNAs were subjected to the enrichment analysis. The enrichment analysis of the up-regulated genes showed that the SARS-CoV-2 down-regulated genes were the top enriched items, while the enrichment analysis of the down-regulated genes showed that both SARS-CoV-2 up- and down-regulated genes were enriched (supplementary figure 3). The compartment enrichment analysis of the up-regulated genes showed that synapse and neuron relevant items were top enriched, while the compartment enrichment analysis of the down-regulated genes showed the extracellular vesicle relevant items were top enriched (supplementary figure 2).

## DISCUSSION

Our results paint the picture of the brain reacting to the inflammatory tidal wave unleashed by COVID-19. Proteomics revealed an almost similar CSF protein profile in COVID-19 patients compared to patients with HSVE, both in patients with and without bacterial superinfection. However, the observed changes were much less pronounced in COVID-19 than in HSVE. Of note, the proteins found to be elevated in COVID-19 were mainly of extrathecal origin as indicated by normal CSF/serum ratios, reflecting probably both the pronounced systemic inflammatory reaction in COVID-19 and the high frequency of minor blood-CSF barrier disruption as previously reported in patients with COVID-19. Moreover, some of the proteins found to be elevated in the CSF, such as LRG1 and the proteins of the Serpin family, are not known to be produced within the CNS but were previously found, including in proteomic studies, in the plasma of patients with COVID-19, further suggesting diffusion from the blood into the CSF *(19, 20)*.

The presence of bacterial superinfection had not been taken into account in previous studies on the neuroimmunological impact of a COVID-19. Bacterial superinfection (BSI) was associated with significantly increased levels of the inflammation-related proteins studied but did not affect the protein pattern itself.

Finally, also the pathway analysis showed a preponderance of immunological pathways being activated in COVID-19_highpct_ patients.

Our study also reaffirms previous studies reporting increased IL-6, CXCL10 and IL-16 serum levels in patients with COVID-19 with BSI being a major bias *(14, 21)*. Results from the array pointed to elevated serum concentrations of integrins such as ICAM1 or hemostasis-related proteins such as SerpinE1 in the serum of COVID-19 patients.

Progranulin levels are significantly increased in the CSF of symptomatic and asymptomatic COVID-19 patients, in patients with and without bacterial superinfection and both in patient with mild and severe symptoms. Progranulin is not known as a marker for neuronal death but for neuroprotection *(22–24)*. One hypothesis for the increase of progranulin is that the brain is reactively increasing its neuroprotective and anti-inflammatory stockpile. Thus, progranulin in the CSF would be a result of microglial and neuronal activation, which has in fact been shown in several neuropathological post-mortem and animal COVID-19 studies *(7–9)*. Progranulin in the rodent brain is most strongly expressed in the olfactory and temporal regions and Sars-CoV2 has been proven to infiltrate this system in the animal model. Another possible explanation could be that progranulin is maybe released during infiltration of these regions *(8, 18, 25)*. The known COVID-19 symptoms dysgeusia and dysosmia are not depending on disease severity. This brings our progranulin data well in line because progranulin levels were also independent from COVID-19 disease severity in our cohort. The CSF/serum ratios are not reasonable here because progranulin is produced also outside the CNS *(26, 27)*. In sum, high progranulin CSF levels and low cytokine CSF/serum ratios together may point at a brain in a defensive state against the inflammatory storm coming from the blood. The specific protein results on their own may explain the normal CSF routine parameter measured here and in other studies *(14, 28, 29)* going along with neurological symptoms as these small concentrations are possibly able to initiate a weak response of the brain.

Normal CSF routine parameters have been seen before also in symptomatic antibody-associated autoimmune encephalitis *(30–32)*. Extensive screening for anti-neural antibodies including on native neuronal tissue did not reveal any pathological titers of known antibodies nor did it reveal any novel, unknown antibody reactivities. Our cohort did comprise some patients with focal neurological symptoms such as oculomotor palsy but most of them had unspecific neurological symptoms such as encephalopathy that might in total lower the risk for antibody detection in our cohort (see Supplementary Table 2). Still, the absence of anti-neural autoantibodies at clinically relevant titers in 32 patients argues against a major role of antibody-mediated mechanisms in the pathophysiology of COVID-19 with neurological symptoms.

Given previous evidence suggesting that SARS-CoV-2 infection of the CNS is very rare, if it occurs at all, it seems unlikely that the altered linear RNA profile in the CSF of patients with COVID-19 and neurological involvement compared to healthy controls and patients with other neurological diseases observed in our study reflects direct SARS-CoV-2-mediated neural damage. Instead, these results show rather indirect, systemic effects in the brain, e.g., diffusion of mediators of inflammation from the peripheral circulation into the CNS, either passively or through a leaky blood-brain barrier, or hypoxia, and/or brain endothelial cell damage. In the enrichment analysis of up-regulated genes in COVID-19 CSF, the significantly enriched items are the down-regulated genes by SARS-CoV-2 in cells or tissues, which indicated that one COVID-19 pathomechanism maybe forces the neurons to secret RNAs and their RNA fragments into the extracellular compartment.

Among the down-regulated linear RNAs in COVID-19 CSF in our study, *HBB (Hemoglobin subunit beta)* was not detectable in every COVID-19 CSF samples, while it was expressed in all other CSF samples. Therefore blood contamination as a source for *HBB* seems to be very unlikely. Maybe this finding is due to the previously reported SARS-CoV-2 attack on the heme on the 1-B chain of hemoglobin eventually causing a lower amount of hemoglobin *(33)*. Disruption of this pathway would lead to a reactive increase of HBB resulting in downregulation. Also, in our enrichment analysis, the down-regulated genes were found to be associated with extracellular vesicles, which suggests that COVID-19 may affect the RNA profile of extracellular vesicles derived from the brain.

## MATERIALS AND METHODS

### Demographic data and clinical features

This study has been approved by the ethics committee of the Charité Universitätsmedzin Berlin (EA176_20). Every patient gave written and informed consent. We performed a retrospective study on CSF and serum samples from patients with PCR-proven SARS-CoV-2 infection in the nasopharyngeal area who underwent lumbar puncture to rule out CNS pathologies such as autoimmune encephalitis or meningitis involving a total of 38 COVID-19 patients. Classification according WHO definition resulted in 24 patients having grade 4 COVID-19, 7 grade 3 COVID-19, 2 grade one or two COVID-19, respectively. Three patients were SARS-CoV-2 positive as part of the general screening without showing any symptoms. Mean age was 68.6 years. 28 were male and 10 female. The indications for performing lumbar puncture were clinical symptoms of encephalopathy (dizziness, delir and headache) or prolonged awakening during the weaning. Specific neurological symptoms were very rare. One patient had myclonia without signs of epilepsy on the EEG and one with oculomotor disturbances after resuscitation. Exclusion criteria were other neuroimmunological diseases such as other virus-meningitis, known history of Multiple Sclerosis or HIV.

We used different group sizes of the control cohort (n=28) and the HSVE cohort due to different sample volumes restrictions. We included 23/28 from the controls without neurological diagnosis from the Charité biobank. The other 5 controls and 10 HSVE patients were recruited from the CSF lab at the university of Magdeburg. These control patients got a spinal tab to exclude neurodegenerative, neuroimmunological diseases, or subarachnoid hemorrhage for what they were not diagnoses in the end. Every herpes simplex virus patient had a positive HSV-PCR test in the CSF at diagnosis. For cytokine measurements CSF and serum from 11 control patients and 10 patients with herpes simplex virus meningoencephalitis were used. Control samples for the linear and circular RNA measurements were recruited from existing cohorts suffering from no neurological disease as negative controls and a positive cohort with multiple sclerosis and Alzheimer’s Disease. COVID-19 patients in the RNA analysis group had symptoms grade 2-4. Normal controls for the Progranulin measurements were also recruited in a age-matched fashion from the existing cohorts from the neurochemical lab at the Department of Neurology, Magdeburg, Germany.

### Cytokine measurements

#### Cytokine array

First, we used an array to analyze cytokine levels in CSF samples of COVID-19 patients (n = 9), HSVE patients (n = 5) and controls (n = 4). We used a semi-quantitative neuroimmunology cytokine array (bio-techne, Minneapolis, MN) according to the manufacturer’s instructions *(15)*. This assay measures 36 human cytokines and inflammation related proteins (CCL1, CCL2, MIP-1a, CCL5, CD40L, C5/C5a, CXCL1, CXCL10, CXCL11, CXCL12, G-CSF, GM-CSF, ICAM-1, IFN-g, IL-1a, IL-1b, IL-1ra, IL-2, IL-4, IL-5, IL-6, IL-8, IL-10, IL-12 p70, IL-13, IL-16, IL-17A, IL-17E, IL-18, IL-21, IL-27, IL-32a, MIF, Serpin E1, TNF-a, TREM-1). We used the software Kodak D1 3.6 (Eastman Kodak, Rochester, New York), for determining the background-corrected sum intensity for each ROI on the membrane. Separate membranes were normalized to each other using the results of the positive controls.

#### ELISA measurements

The levels of IL-6, IL-16 and CXCL10 were measured in CSF samples and sera of 18 COVID-19 patients with mild to severe symptoms (12 COVID-19_lowPCT_ and 6 COVID-19_highPCT_), 10 HSVE patients and 11 normal controls using commercially available specific human Quantikine ELISA (bio-techne, Minneapolis, MN). The assays were performed following the instructions provided by the manufacturer.

#### Progranulin measurements

We measured Progranulin in 21 COVID-19 patients (mean age 71.7yrs, 6 female 17 male) with 12 belonging to the C19_lowPCT_ cohort and 9 to the C19_highPCT_ cohort. Progranulin is differently metabolized in the CNS than outside the CNS and we therefore did not measured blood levels *(26, 27, 34)*. Again, we used non-inflammatory, non-neurodegenerative controls (n=22) but now from the CSF lab at the Department of Neurology, University Hospital Magdeburg, Germany. Progranulin measurements were done as describe before with a commercial ELISA *(25, 35)*.

#### Autoantibody measurements

Thirty-two CSF and 32 serum samples from the same time from 32 COVID-19 patients were tested for anti-neuronal and anti-glial autoantibodies.Testing was performed by Laboratory Prof. Dr. Stöcker, Lübeck, Germany (N=15),and Labor Berlin, Berlin, Germany (N=19). Two patients were measured in both laboratories, one with Yo-ab and one with NMDAR-ab. At Labor Berlin antibody screening in CSF/serum included testing for antibodies against NMDAR, LGI-1, Caspr2, DPPX, AMPAR, GABAbR, aquaporin-4, myelin, glycine receptor, dopamin-2R, mGluR5, GAD-65 using cell-based assays (CBA) and 12 paraneoplastic antibodies (anti-CV2, - Hu, -Ri, -Yo, -Ma2/Ta, -Zic4, -titin, -SOX1, -amphiphysin, -GAD65, -recoverin, Tr/DNER using immunblots (Euroimmun, Germany). At the Laboratory Prof. Dr. Stöcker the same panel of 24 antibodies tested by Labor Berlin plus 12 additional antibodies (anti-CARPVIII, mGluR1, - GABAaR, -ARHGAP26/anti-Ca, ITPR1/anti-Sj, -Homer3, -Neurexin-3 alpha, -MOG, - neurochondrin, -IGLON-5, -flotillin-1 and -2) were measured by CBA. In addition, all serum and CSF samples at the Laboratory Prof Dr. Stöcker were tested for novel autoantibody reactivities by immunohistochemistry on native hippocampus and cerebellum tissue sections. Every panel, in every laboratory included antibodies to extracellular, synaptic, and intracellular epitopes.

### Mass spectrometry

#### Sample preparation

Cerebrospinal fluid (CSF) samples were reduced, alkylated, digested and conditioned on a Biomek i7 workstation, as previously described *(19)*. Volumes and concentration of reagents were adapted to accommodate the increased input volume. In short, 50 µl CSF was transferred to 170 µl 8M Urea, 5 mM DTT, 100 mM ABC and incubated at 30C for 1 h before addition of 18 µl 120 mM IAA and incubated for 30 min at RT in the dark. 450 µl of 100 mM ABC buffer were added, and 688 µl of the solution digested with 1.25 ug trypsin in a total volume of 700 µl and approx. 2 M urea, at 37°C ON. The reaction was stopped by addition of 28 µl 25% formic acid. 725 µl of peptides were cleaned by solid phase extraction, vacuum dried, reconstituted in 50 µl 0.1% formic acid and their concentration determined (Pierce Quantitative Fluorometric Peptide Assay (number 23290)).

#### Mass spectrometry

Approximately five micrograms of peptides (5 µl) were used per injection for the 5 min gradient scanning Swath LC-MS/MS *(36)*. In brief, the peptides were resolved on an Agilent 1290 Infinity II system coupled to a TripleTOF 6600 mass spectrometer (SCIEX) equipped with IonDrive Tubo V Source (Sciex). For reverse phase separation we used a C18 ZORBAX Rapid Resolution High Definition (RRHD) column (2.1mm × 50mm, 1.8µm particles) at a flow rate of 0.8 ml/min and column temperature of 30°C. A linear gradient was applied which ramps from 1% B to 40% B in 5 minutes (Buffer A: 0.1% FA; Buffer B: ACN/0.1% FA) with a flow rate of 800 µl/min. For washing the column, the organic solvent was increased to 80% B in 0.5 minutes and was kept for 0.2 minutes at this composition before going back to 1% B in 0.1 min. The column was equilibrated for 2.1 min before the next injection. The mass spectrometer was operated in high sensitivity mode.

The data was recorded using scanning SWATH with a precursor isolation window of 10 m/z and a mass range of 400–900 m/z that was covered in 0.5 s. Nebulizer gas temperature, heater gas and curtain gas were set to 50, 40 and 25, respectively. The source temperature was set to 450 and the ion spray voltage to 5500V. Raw data were binned in the quadrupole or precursor dimension into 8-m/z bins.

#### Data processing

The raw data were processed using DIA-NN 1.7.12 *(37)* as previously described *(19)*. MS2 and MS1 mass accuracies were set to 20 and 12 ppm, respectively, and scan window size set to 6. A spectral library was generated from a published list of peptides present in human CSF (PXD015087, 8250 peptides) *(38)* using the Deep learning-based spectra and RT prediction provided in DIA-NN. The precursors were annotated using the Human UniProt sequence database (Human_UP000005640_9606, accessed 2019-12-20) *(39)*. The library was automatically refined based on the dataset in question at 0.01 global q-value (using the “Generate spectral library” option in DIA-NN).

Obtained after pre-treatment and annotation with DIA-NN, peptide intensities of each sample were subjected to quality control. Two samples with low quality were removed.Weighted mean scaling of intensities (with a weight proportional to square of peptide presence and inversely proportional to its standard deviation) was applied to align sample intensity distributions within each group. Peptides with excessive missing values (> 35% per group) were excluded from analysis. To the rest of peptides group-based imputation of missing values using PCA method *(40)* was applied. Then they were normalized using LIMMA *(41)* implementation of cyclicloess method *(42)* with option “fast” *(43)*. To obtain a quantitative protein data matrix, the log2-intensities of peptides were filtered, only peptides belonging to one protein group were kept, and then summarised by “maxLFQ” method (*(44)*, implemented in R package iq *(45)*) into protein log intensity. Median scaling of protein intensities was applied. Change of the data matrix after selected pre-processing steps is presented in supplementary table 3. Description of sample groups along with some demographic information is provided in supplementary table 4.

#### Data analysis

The goals were 1) to find similarities/dissimilarities in the response to COVID-19 and to HSVE Meningitis and 2) to reveal the influence of bacterial superinfection on the neuroimmunological response. Statistical analysis of proteomics data was carried out using R scripts based on publicly available packages. Linear modelling was based on the R package LIMMA *(41)*. Two models were applied to the data set.

Model 1: log_2_(*p*) ∼ 0+Class. Here, log_2_(*p*) is log2 transformed expression of a protein, the categorical factor Class has four levels: control, C19_low, C19_high, HSVE; reference level is control. The contrasts evaluated within Model 1 are listed in table 2.

For finding regulated features following criteria were applied:

Significance level alpha was set to guarantee the false discovery rate below ∼ 5% across all contrasts. We found that *a* = 0.01 was delivering the required level of Benjamini–Hochberg FDR *(46)*.

The log fold change criterion was applied to guarantee that the measured signal is above the average noise level. As such, we have taken median residual standard deviation of the model: log_2_(T) = median residual SD of linear modelling.

Another set of selection criteria was applied for creation of heatmaps. We applied very strict criteria: *a* = 0.001 was delivering BHFDR below 1% and log_2_(T) = 1.5*median residual SD

Model 1 allowed to address the differences between clinical groups. Note that control and HSVE groups were confounded with gender and age (see Tab Change matrix) and this limits the accuracy of our findings.

Model 2: log_2_(*p*) ∼ 0+sex + f2 + f3.

The categorical factor sex has two levels: female and male; reference level is male, factor f2 takes into account PCT level and f3 differences in the WHO grade.

Model 2 was devoted to more accurate evaluation of the influence of bacterial superinfection on the proteome of COVID patients. For this we used transformed numerical values of PCT levels f2 = log_2_(1+*PCT*). As COVID-19 groups had roughly the same proportion of females (∼0.2), sex was introduced into the model as a categorical factor. COVID-19 patients were of different WHO grade, which was confounded with PCT levels, therefore, to minimize its influence on the effect of BSI it was included into the model as a numerical factor f3. Exploratory analysis revealed that effect of age was relatively small and therefore was skipped from the analysis.

For model 2 the significance level alpha was set to guarantee the false discovery rate below ∼ 10% for contrast f2. We found that alpha = 0.01 was delivering required level of BH FDR. The log fold change threshold was set as 0.5.

#### Functional analysis of proteomics data

Functional analysis (GSEA) was carried out using R package clusterProfiler (Yu 2012). For selecting the most (de)regulated GO terms we applied filter: 2 ≤ term size ≤ 250. Analyses were carried out with Benjamini–Hochberg FDR threshold 0.3.

### The RNA sequencing of CSF and data analysis

#### Epidemiology of the circRNA measurements

In the RNA sequencing, we used the CSF of 5 COVID-19 patients and 4 CSF samples from the control cohort. For comparison with other neurological diseases, we used an already established RNA data set of neurological diseases cohort (nd-cohort) at the Kjems’ lab at the Department of Molecular Biology and Genetics at Aarhus University, Denmark. This nd cohort comprises 4 patients with multiple sclerosis at diagnosis and 4 with Alzheimer’s disease.

#### CSF RNA purification and genomic DNA removal

CSF samples were thawed in cold room (4°C) and then centrifuged at 5000 g 4 °C for 10min. The supernatant was taken out for the RNA purification.

For total RNA sequencing, 1ml CSF was used for RNA purification using miRNeasy Serum/Plasma advanced kit (QIAGEN) according to the manufacturer’s protocol and the RNA was eluted in 50 µl RNase-free water. The genomic DNA removal was done by using Turbo DNA-free kit (Ambion) according to the manufacturer’s instructions and 50 µl DNA-free RNA was taken out. The RNA was concentrated by adding 50 µl RNase-free water, 10 µl 3 M, pH 5.5 sodium acetate, 250 µl pre-chilled 99% ethanol and 1 µl Glycoblue (Ambion, MA, USA), and incubating at −20 °C overnight. RNA was pelleted by centrifugation at 16,000 *g* for 20 min at 4 °C and the pellet was washed using 1ml 80% ethanol. The RNA was re-pelleted by centrifugation at 10,000 g for 5min and the RNA pellet was re-suspended in 12 µl RNase free water. The RNA was stored in -80 °C.

#### Total RNA sequencing

The total RNA sequencing library was constructed by using 8µl of the RNA purified from 1 ml CSF and SMARTer Stranded Total RNA-Seq Kit v2 - Pico Input Mammalian (Takara). The fragmentation time was 90 s and the rest of the experiment followed the protocol designed for the 250 pg RNA input. The Bioanalyzer High sensitivity DNA analysis kit (Agilent) was used to determine the size of the library fragments and the KAPA Library Quantification kit (Roche) was used to quantify the library. The libraries were pooled with equal amount and sequenced on a Novaseq sequencing machine using SP flowcell paired end 100 cycles (Illumina).

#### Total RNA sequencing data analysis

Sequencing data were pre-processed by removing adapter sequence and trimming away low quality bases with a Phred score below 20 using Trim Galore (v0.4.1). Quality control was performed using FastQC to ensure high quality data. Quantification of gene expression was performed by mapping the filtered reads to the human genome (hg19) using Tophat2. The software featureCounts was used to quantify the number of reads mapping to each gene using gene annotation from the Gencode release 29. The linear RNAs were included in the further analysis if they had expression in all the samples of at least one group (COVID-19 group, Healthy group or Neurological disease group).Quantification of circRNA was done primarily using CIRI2 and secondarily using find_circ to support circRNA findings. Using bedtools detected circRNAs were checked against identical circRNAs annotated in circBase (PMID: 25234927), CIRCpedia (PMID: 30172046) and circAtlas (PMID: 32345360).

Differential expression analysis was performed using DESeq2 in R on the combined gene and circRNA expression levels. Volcano plots, dot plots, PCA plots were done in R. The linear RNA and circRNAs were considered to be significantly differential expressed with the FDR < 0.05.

The enrichment analysis was done using Enrichr https://maayanlab.cloud/Enrichr/)*(47)*

## Data Availability

All data produced in the present study are available upon reasonable request to the authors

## List of Supplementary Materials

Supplementary figures :3

Supplementary tables :4

## Acknowledgments

The authors are grateful to Kerstin Kaiser und Jeanette Witzke, Neurochemistry laboratory, Department of Neurology, University hospital Magdeburg, Magdeburg, Germany

## Funding

### Author contributions

Conceptualization: DR,VF,YY,KL,HR,JK,MM,FH,PK

Methodology: DR,KG,AG,Vf,MR,MM,YY,MV,JK,SS,HR,FH,PK

Investigation:DR,KG,VF,MM,YY,MV,JK,SS,HR,CO,KR,PS,FK,CM,HS,JK,SV,FHP,BT,WS,P K

Visualization: DR,KG,AG,VF,MM,YY,VM,PK Funding acquisition: HR,ED,FH,JK,MR

Project administration: HR, CM,HS,JK,FHP,PK

Supervision: FH,MR,LES,PK

Writing – original draft: PK,DR,VF,YY,MM

Writing – review & editing: PK, DR,VF,YY,MM,ME,SJ,KR,SS, JK,

### Data and materials availability

All data, code, and materials used in the analysis are available per email request to the corresponding author

## Abbreviations

AD: Alzheimer’s dementia
ARHGAP26: Rho GTPase-Activatin Protein 26
AMPAR: -Amino-3-hydroxy-5 methyl-4-isoazolepropionic acid receptor
BSI: Bacterial Superinfection
Caspr2: Contactin-associated protein 2
CBA: Culture based assay
CNS: Central Nervous System
COVID-19: Coronavirus Disease 2019
CSF: Cerebrospinal fluid
CV2/CRMP5: Cytoplasmatic protein 5
CXCL-10: C-X-C motif chemokine 10
DPPX: Dipeptidyl-Peptidase-like Protein-6
EEG: Electroencephalogramm
ELISA: Enzyme-linkend Immunfluorescene Assay
GABAaR: Gamma-amino-butyrat-a-receptor
GABAbR: Gamma-amino-butyrat-b-receptor
GAD-65: Glutamic Acid Decarboxylase 65
HSVE: Herpes-simplex virus meningoencephalitis
IL-6: Interleukine- 6
LGI-1: Leucin-rich glioma inactivated Protein 1
mGluR1: metabotropic Glutamat Receptor 1
mGluR5: metabotropic Glutamta-receptor 5
MOG: Myelin-Oligodendrocyte Glycoprotein
NMDAR: N-Methyl-D-Aspartate Receptor
PCT: Procalcitonin
PGRN: Progranulin
SARS-CoV2: Severe Acute Respiratory Syndrome Coronavirus 2
Tr/DNER: Delta/notch-like epidermal growth factor
ITPR1/anti-Sj: Inositol 1,4,5-trisphosphate receptor type 1

