## Supplemental Data 1 for "The Brain Reacting to COVID-19: Analysis of the Cerebrospinal Fluid and Serum Proteome,Transcriptome and Inflammatory Proteins"

### Supplementary material

#### Supplementary Table 1: Protein Abbreviation List for Figure 1

| Protein.Names | First.Protein.Description |
|---|---|
| VGF_HUMAN | Neurosecretory protein VGF |
| TYRO3_HUMAN | Tyrosine-protein kinase receptor TYRO3 |
| TIMP2_HUMAN | Metalloproteinase inhibitor 2 |
| TIMP1_HUMAN | Metalloproteinase inhibitor 1 |
| TICN1_HUMAN | Testican-1 |
| SPRL1_HUMAN | SPARC-like protein 1 |
| SPRC_HUMAN | SPARC |
| SLIK1_HUMAN | SLIT and NTRK-like protein 1 |
| SHPS1_HUMAN | Tyrosine-protein phosphatase non-receptor type substrate 1 |
| SGCE_HUMAN | Epsilon-sarcoglycan |
| SE6L2_HUMAN | Seizure 6-like protein 2 |
| IC1_HUMAN | Plasma protease C1 inhibitor |
| A2AP_HUMAN | Alpha-2-antiplasmin |
| AACT_HUMAN | Alpha-1-antichymotrypsin |
| A1AT_HUMAN | Alpha-1-antitrypsin |
| SEM7A_HUMAN | Semaphorin-7A |
| LYAM1_HUMAN | L-selectin |
| 7B2_HUMAN | Neuroendocrine protein 7B2 |
| SCG2_HUMAN | Secretogranin-2 |
| RNAS4_HUMAN | Ribonuclease 4 |
| RNAS1_HUMAN | Ribonuclease pancreatic |
| RGMB_HUMAN | RGM domain family member B |
| RET4_HUMAN | Retinol-binding protein 4 |
| EPCR_HUMAN | Endothelial protein C receptor |
| PRIO_HUMAN | Major prion protein |
| KPYM_HUMAN | Pyruvate kinase PKM |
| PGRP2_HUMAN | N-acetylmuramoyl-L-alanine amidase |
| PEBP4_HUMAN | Phosphatidylethanolamine-binding protein 4 |
| PCSK1_HUMAN | ProSAAS |
| A1AG2_HUMAN | Alpha-1-acid glycoprotein 2 |
| A1AG1_HUMAN | Alpha-1-acid glycoprotein 1 |
| OMGP_HUMAN | Oligodendrocyte-myelin glycoprotein |
| NRX2A_HUMAN | Neurexin-2 |
| NRCAM_HUMAN | Neuronal cell adhesion molecule |
| NPTXR_HUMAN | Neuronal pentraxin receptor |
| NPTX1_HUMAN | Neuronal pentraxin-1 |
| NEGR1_HUMAN | Neuronal growth regulator 1 |
| LYSC_HUMAN | Lysozyme C |
| LYVE1_HUMAN | Lymphatic vessel endothelial hyaluronic acid receptor 1 |
| LYNX1_HUMAN | Ly-6/neurotoxin-like protein 1 |
| LY6H_HUMAN | Lymphocyte antigen 6H |
| A2GL_HUMAN | Leucine-rich alpha-2-glycoprotein |
| LAMP2_HUMAN | Lysosome-associated membrane glycoprotein 2 |
| K2C1_HUMAN | Keratin, type II cytoskeletal 1 |
| KLKB1_HUMAN | Plasma kallikrein |

| | |
|---|---|
| KLK6_HUMAN | Kallikrein-6 |
| IGJ_HUMAN | Immunoglobulin J chain |
| ITIH4_HUMAN | Inter-alpha-trypsin inhibitor heavy chain H4 |
| ITIH2_HUMAN | Inter-alpha-trypsin inhibitor heavy chain H2 |
| ITIH1_HUMAN | Inter-alpha-trypsin inhibitor heavy chain H1 |
| LV319_HUMAN | Immunoglobulin lambda variable 3-19 |
| LV147_HUMAN | Immunoglobulin lambda variable 1-47 |
| IGLL5_HUMAN | Immunoglobulin lambda-like polypeptide 5 |
| IGLC3_HUMAN | Immunoglobulin lambda constant 3 |
| KV401_HUMAN | Immunoglobulin kappa variable 4-1 |
| KV320_HUMAN | Immunoglobulin kappa variable 3-20 |
| KV105_HUMAN | Immunoglobulin kappa variable 1-5 |
| IGKC_HUMAN | Immunoglobulin kappa constant |
| S4R460_HUMAN | Immunoglobulin heavy variable 3/OR16-9 (non-functional) |
| IGHM_HUMAN | Immunoglobulin heavy constant mu |
| IGHG3_HUMAN | Immunoglobulin heavy constant gamma 3 |
| IGHG1_HUMAN | Immunoglobulin heavy constant gamma 1 |
| IGHA2_HUMAN | Immunoglobulin heavy constant alpha 2 |
| IGHA1_HUMAN | Immunoglobulin heavy constant alpha 1 |
| IBP7_HUMAN | Insulin-like growth factor-binding protein 7 |
| IBP6_HUMAN | Insulin-like growth factor-binding protein 6 |
| IBP4_HUMAN | Insulin-like growth factor-binding protein 4 |
| IBP2_HUMAN | Insulin-like growth factor-binding protein 2 |
| PGBM_HUMAN | Basement membrane-specific heparan sulfate proteoglycan core protein |
| SAP3_HUMAN | Ganglioside GM2 activator |
| FIBG_HUMAN | Fibrinogen gamma chain |
| FIBB_HUMAN | Fibrinogen beta chain |
| FIBA_HUMAN | Fibrinogen alpha chain |
| FCGBP_HUMAN | IgGFc-binding protein |
| FA5_HUMAN | Coagulation factor V |
| CATD_HUMAN | Cathepsin D |
| CATB_HUMAN | Cathepsin B |
| CBPQ_HUMAN | Carboxypeptidase Q |
| CBPE_HUMAN | Carboxypeptidase E |
| CERU_HUMAN | Ceruloplasmin |
| CNTN2_HUMAN | Contactin-2 |
| CLUS_HUMAN | Clusterin |
| CSTN3_HUMAN | Calsyntenin-3 |
| CSTN1_HUMAN | Calsyntenin-1 |
| SCG1_HUMAN | Secretogranin-1 |
| CFAI_HUMAN | Complement factor I |
| CFAD_HUMAN | Complement factor D |
| CFAB_HUMAN | Complement factor B |
| CADH2_HUMAN | Cadherin-2 |
| CD14_HUMAN | Monocyte differentiation antigen CD14 |
| CADM2_HUMAN | Cell adhesion molecule 2 |
| CA2D1_HUMAN | Voltage-dependent calcium channel subunit alpha-2/delta-1 |
| CO9_HUMAN | Complement component C9 |
| CO7_HUMAN | Complement component C7 |
| CO6_HUMAN | Complement component C6 |

| | |
|---|---|
| CO5_HUMAN | Complement C5 |
| CO4A_HUMAN | Complement C4-A |
| CO2_HUMAN | Complement C2 |
| C1R_HUMAN | Complement C1r subcomponent |
| C1QC_HUMAN | Complement C1q subcomponent subunit C |
| C1QB_HUMAN | Complement C1q subcomponent subunit B |
| B4GA1_HUMAN | Beta-1,4-glucuronyltransferase 1 |
| B2MG_HUMAN | Beta-2-microglobulin |
| ZA2G_HUMAN | Zinc-alpha-2-glycoprotein |
| VAS1_HUMAN | V-type proton ATPase subunit S1 |
| A4_HUMAN | Amyloid-beta A4 protein |
| APOC1_HUMAN | Apolipoprotein C-I |
| APOA4_HUMAN | Apolipoprotein A-IV |
| APOA2_HUMAN | Apolipoprotein A-II |
| APOA1_HUMAN | Apolipoprotein A-I |
| APLP2_HUMAN | Amyloid-like protein 2 |
| APLP1_HUMAN | Amyloid-like protein 1 |
| AMBP_HUMAN | Protein AMBP |
| ADA22_HUMAN | Disintegrin and metalloproteinase domain-containing protein 22 |
| A2MG_HUMAN | Alpha-2-macroglobulin |

**Supplementary Table 2: Serum autoantibody findings and associated clinical characteristics in 10 patients with COVID-19.**

| **Patient** | **Antibody** | **Syndrome** |
|---|---|---|
| #1 | NMDAR 1:100 | Nystagmus, orofacial myoclonus (first present after resuscitation),<br>MRI:hypoxic brain damage |
| #2 | Yo 1:100 | Brachiofacial myclonia, EEG normal |
| #3 | Caspr2 1:32 | Unsteady gait and delir |
| #4 | Myelin 1:100 | Protracted waking after mechanical ventilation |
| #5 | Myelin 1:100 | Hyperactive delir |
| #6 | Myelin 1:100 | Delir |
| #7 | Myelin 1:100 | Delir |
| #8 | Caspr2 1:10 and | Very severe delir |

| | | |
|---|---|---|
| | NMDAR 1:10 | |
| #9 | Glycin 1:10 | Mnestic difficulties (known AD) |
| #10 | Glycin 1:10 | Myalgia, delir |

**Supplementary Table 3. Change of the data matrix size** after pre-processing steps.

| Step | N features | M samples |
|---|---|---|
| Input data matrix size | 5486 | 74 |
| After sample removal | 5486 | 72 |
| After features with NA>35% removal | 2230 | 72 |
| After filtering out nonproteotypic peptides | 1992 | 72 |
| After peptides summarization | 271 | 72 |
| Final protein matrix size | 271 | 72 |

**Supplementary Table 4. Clinical groups used for analysis along with some demographic information.** COVID-19 Patients were divided into cohort with elevated Procalcitonin levels (PCT>1) indicating a confounding superinfection (C19_high_PCT) and a cohort C19_low_PCT with normal Procalcitonin levels (PCT ≤ 1). All C19_high_PCT patients had WHO severity grade 4, C19_low_PCT patients on average had WHO severity grade 3. Two other groups were control and HSV Meningitis (HSVE).

| | **Female** | | | **Male** | | | **Total** | | |
|---|---|---|---|---|---|---|---|---|---|
| | **Avg Age** | **Avg PCT** | **NN pat** | **Avg Age** | **Avg PCT** | **NN pat** | **Avg Age** | **Avg PCT** | **NN pat** |

| | | | | | | | | |
|---|---|---|---|---|---|---|---|---|
| C19_high_PCT | 63.7 | 1.8 | 3 | 71.8 | 4.0 | 13 | 70.3 | 3.6 | 16 |
| C19_low_PCT | 69.2 | 0.1 | 5 | 68.8 | 0.3 | 17 | 68.9 | 0.2 | 22 |
| control | 57.6 | | 17 | 58.8 | | 11 | 58.1 | | 28 |
| HSVE | 53.2 | | 6 | | | | 53.2 | | 6 |
| **Total** | **59.2** | | **31** | **67.0** | | **41** | **63.7** | | **72** |

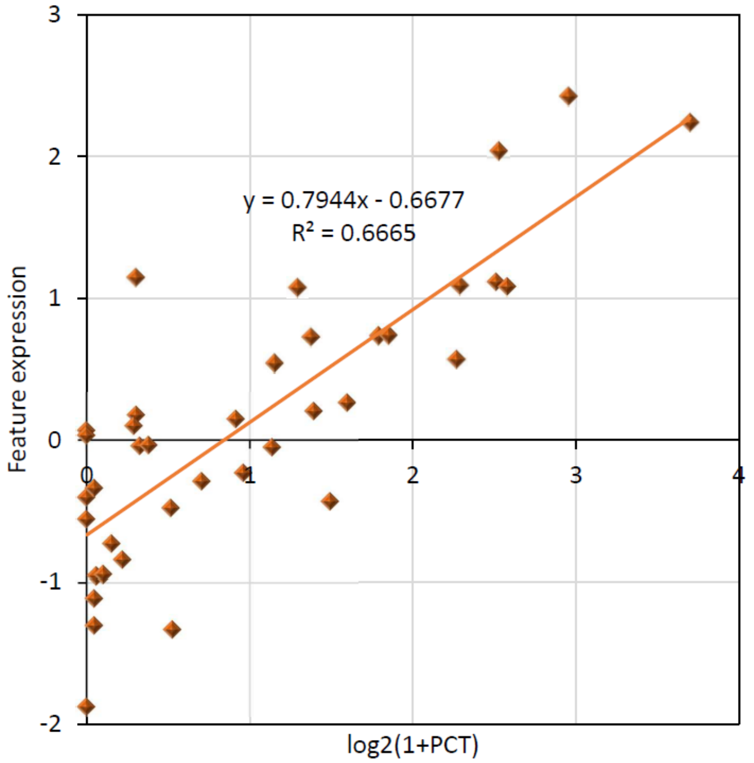


Supplementary Figure 1. Linear dependence of combined log2 expression of three proteins y= (C4A + CD14)/2 – NRCAM on transformed PCT level x = log2(1 + PCT). Proteins C4A and CD14 show increase of expression with increase of x and have nearly opposite dependence on sex and WHO grade, while NRCAM show decrease with increase of x and is almost independent on sex and WHO grade. Linear fit showed that approximately 67% of the variation in combined proteins expression can be explained by BSI ($R^2$ = 0.67). The fit was conducted on all COVID samples (as shown in the Figure) and on WHO grade 4 only samples with practically the same outcome.

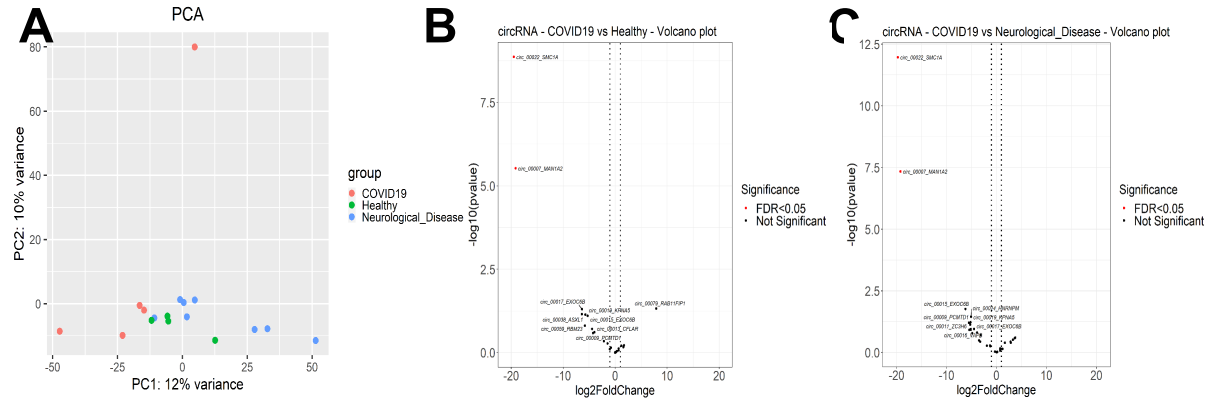


Supplementary Figure 2. CircRNA data analysis. (A) Principal component analysis (PCA) of circRNA profile in CSF of Covid-19 patients, healthy controls and patients with neurological disease. Differential expression analysis of circRNAs in (B) CSF of Covid-19 patients and healthy controls and (C) CSF of Covid-19 patients and patients with neurological disease. Red dots: FDR < 0.05.

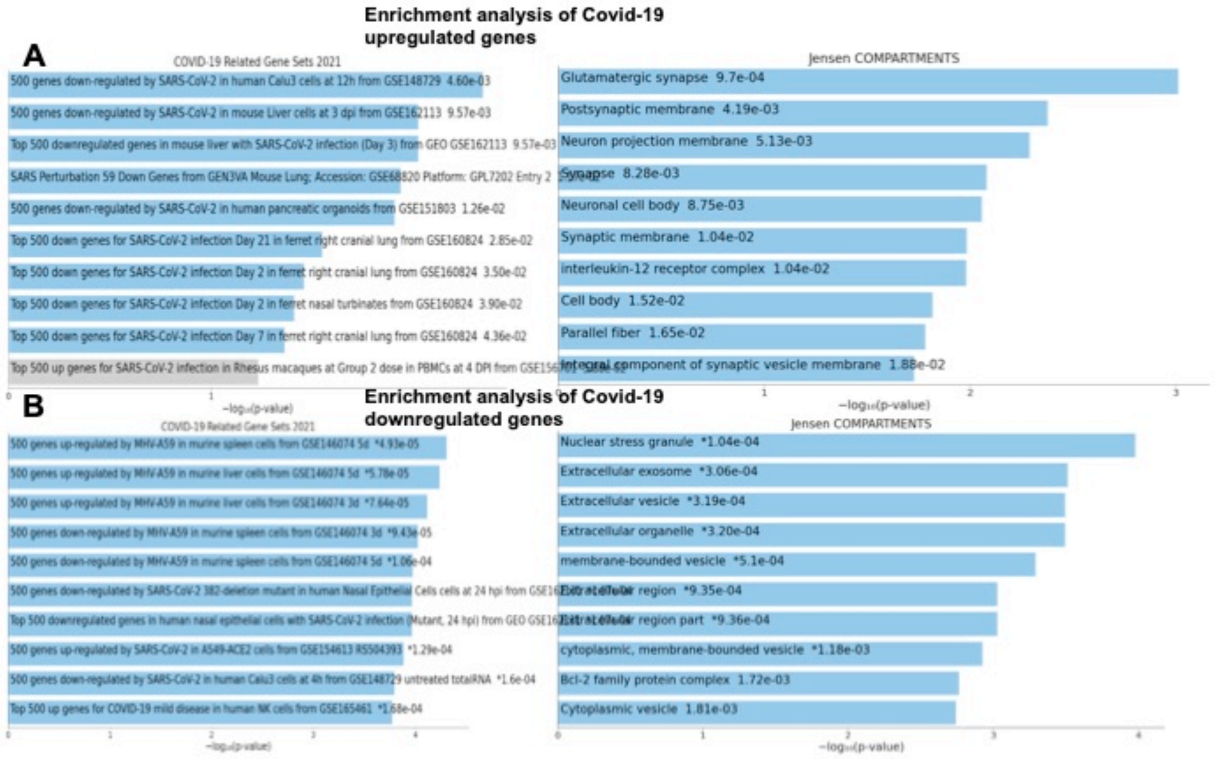


Supplementary Figure 3 Supplementary Figure. Enrichment analysis of deregulated genes in COVID-19 CSF. Top 10 enriched items in the COVID-19 related gene sets 2021 and cellular compartment localization in the analysis of (A) COVID-19 up-regulated genes and (B) COVID-19 down-regulated genes. The p-value is listed after each item; the items with p-value < 0.05 are significantly enriched; An asterisk (*) next to a p-value indicates the term also has a adjusted p-value <0.05.
